# Artificial Intelligence and Machine Learning in Prehospital Emergency Care: A Systematic Scoping Review

**DOI:** 10.1101/2023.04.25.23289087

**Authors:** Marcel Lucas Chee, Mark Leonard Chee, Haotian Huang, Katie Mazzochi, Kieran Taylor, Han Wang, Mengling Feng, Andrew Fu Wah Ho, Fahad Javaid Siddiqui, Marcus Eng Hock Ong, Nan Liu

## Abstract

**Introduction:** The literature on the use of AI in prehospital emergency care (PEC) settings is scattered and diverse, making it difficult to understand the current state of the field. In this scoping review, we aim to provide a descriptive analysis of the current literature and to visualise and identify knowledge and methodological gaps using an evidence map.

**Methods:** We conducted a scoping review from inception until 14 December 2021 on MEDLINE, Embase, Scopus, IEEE Xplore, ACM Digital Library, and Cochrane Central Register of Controlled Trials (CENTRAL). We included peer-reviewed, original studies that applied AI to prehospital data, including applications for cardiopulmonary resuscitation (CPR), automated external defibrillation (AED), out-of-hospital cardiac arrest, and emergency medical service (EMS) infrastructure like stations and ambulances.

**Results:** The search yielded 4350 articles, of which 106 met the inclusion criteria. Most studies were retrospective (n=88, 83·0%), with only one (0·9%) randomised controlled trial. Studies were mostly internally validated (n=96, 90·6%), and only ten studies (9·4%) reported on calibration metrics. While the most studied AI applications were Triage/Prognostication (n=52, 49·1%) and CPR/AED optimisation (n=26, 24·5%), a few studies reported unique use cases of AI such as patient-trial matching for research and Internet-of-Things (IoT) wearables for continuous monitoring. Out of 49 studies that identified a comparator, 39 reported AI performance superior to either clinicians or non-AI status quo algorithms. The minority of studies utilised multimodal inputs (n=37, 34·9%), with few models using text (n=8), audio (n=5), images (n=1), or videos (n=0) as inputs.

**Conclusion:** AI in PEC is a growing field and several promising use cases have been reported, including prognostication, demand prediction, resource optimisation, and IoT continuous monitoring systems. Prospective, externally validated studies are needed before applications can progress beyond the proof-of-concept stage to real-world clinical settings.

**Funding:** This work was supported by the Duke-NUS Signature Research Programme funded by the Ministry of Health, Singapore.

**Research in context:** 

**Evidence before the study:** There has been growing research into artificial intelligence as a potential decision support tool in prehospital emergency care (PEC) settings. Previous reviews summarising AI research in emergency and critical care settings exist, some of which include prehospital care studies peripherally. However, the landscape of AI research in PEC has not been well characterised by any previous review. In this scoping review, we search six databases up to 14 December 2021 for eligible studies and summarise the evidence from 106 studies investigating AI applications in PEC settings.

**Added value of the study:** To our knowledge, our scoping review is the first to present a comprehensive analysis of the landscape of AI applications in PEC. It contributes to the field by highlighting the most studied AI applications and identifying the most common methodological approaches across 106 included studies. Our study examines the level of validation and comparative performance of AI application against clinicians or non-AI algorithms, which offers insight into the current efficacy of AI in PEC. We provide a unique contribution by visualising knowledge and methodological gaps in the field using an evidence map. This scoping review is a valuable resource for researchers and clinicians interested in the potential of AI in PEC and serves as a roadmap for future research.

**Implications of all the available evidence:** Our findings reveal a promising future for AI in PEC, with many unique use cases and applications already showing good performance in internally validated studies. However, there is a need for more rigorous, prospective validation of AI applications before they can be implemented in clinical settings. This underscores the importance of explainable AI, which can improve clinicians’ trust in AI systems and encourage the validation of AI models in real-world settings.

## Introduction

Artificial intelligence (AI) and machine learning (ML) are at the forefront in the era of digital medicine (1, 2). They have been extensively applied to various medial domains such as cardiology (3), ophthalmology (4), emergency medicine (5, 6), and many others. As summarized in numerous reviews and discussions on the adoption of AI and ML techniques in healthcare, both structed and unstructured data (medical images, clinical free texts, time-series physiological signals) benefit from the versatility and flexibility of AI and ML techniques. In addition to healthcare institution-based applications, the intersection of the Internet-of-Things (IoT) and AI have also attracted interest in the form of wearables and remote continuous health monitoring (7).

While there have been attempts to summarise the evidence on AI and ML applications in acute care (5, 6, 8-11), little is reported on their use in prehospital emergency care (PEC) setting. Adoption of AI solutions in PEC is hindered by limited resources and the fast-paced nature of PEC workflows. PEC systems are further complicated by the need for coordination and collaboration between multiple disciplines, such as emergency medicine, critical care, disaster management, and transportation networks. Despite growing research into AI and ML in PEC, there is no systematic review and summary of relevant literature, making it difficult to understand the current state and future directions for the field.

In this paper, we present a systematic scoping review of six databases (MEDLINE, Embase, Scopus, IEEE Xplore, ACM Digital Library, and Cochrane Central Register of Controlled Trials (CENTRAL)) to summarize the current literature on AI and ML applications in PEC research. The aims of the study are to provide a descriptive analysis of the current literature, and to visualise and identify knowledge and methodological gaps using an evidence map (12, 13). The evidence map categorises studies by both applications and input data, allowing a granular analysis of gaps in the current literature.

## Methods

We reported this systematic review according to the Preferred Reporting Items for Systematic Reviews and Meta-Analyses Extension for Scoping Reviews (PRISMA-ScR) checklist (Supplementary File S1). A review protocol was developed but was not publicly registered.

### Literature search and selection criteria

We performed a systematic literature search in six databases, namely, PubMed, Embase, Scopus, IEEE Xplore, ACM Digital Library, and CENTRAL from inception to 14 December 2021. We selected PubMed, Embase, and Scopus for their broad coverage of biomedical and general scientific literature, IEEE Xplore and ACM Digital library to capture more specialised research on AI, and CENTRAL for its focus on controlled trials. We combined two broad concept sets on AI and PEC to conduct our search. A truncated search strategy listing the first three keywords in each set is shown here: (“Artificial intelligence” OR “Deep learning” OR “Machine learning” OR …) AND (“emergency medical service” OR “emergency health service” OR “prehospital” OR …). The full search strategy can be found in Supplementary File 2.

We included original articles that applied AI to PEC data. In this review, we considered articles to have applied AI if they used any of the following AI models: random forest, support vector machine, K-nearest neighbours, neural networks (including deep learning), gradient boosted machine, classification and regression tree, clustering, or natural language processing. We defined PEC to include applications for cardiopulmonary resuscitation (CPR) and automated external defibrillators (AEDs), out-of-hospital cardiac arrests (OHCA), and ambulances or emergency medical service (EMS) stations, but excluded applications in disaster and military medicine. Articles were excluded if they were duplicated, abstracts, or reviews. No restrictions on language were imposed; MLC1 is fluent in Mandarin Chinese and articles in other languages were translated using Google Translate, if necessary.

### Literature selection and data extraction

We exported all extracted literature entries into Microsoft Excel (Office 365) for screening and selection. Each article was independently screened by title and abstract initially, and then full-text by two of three reviewers (MLC1, KM, KT). Discrepancies were resolved through discussions among the two reviewers until consensus was achieved. There was substantial inter-rater agreement, with 96·2% absolute agreement and Cohen’s kappa statistic=0·629. Subsequently, MLC1, MLC2, and HH conducted information extraction from the included literature and all authors reviewed the results. We retrieved information from full-text articles of all included studies, including publication year, study aims, country of dataset origin, AI methods used, comparators used and performance of AI against comparators, study design, sample size and outcomes of interests in predictive modelling studies, input types used, and a summary of each study. We also recorded the study type according to the Transparent reporting of a multivariable prediction model for individual prognosis or diagnosis (TRIPOD) classification of predictive models (14). The TRIPOD classification describes whether a study conducted model development, model validation, or both, as well as the type of model validation, if applicable.

### Evidence map analysis

To investigate the knowledge gap in the current literature, we conducted an evidence map analysis of selected studies. We categorized the studies into one of the following applications: “CPR/AED optimisation”, “Triage/Prognostication”, “ECG interpretation” (electrocardiogram interpretation), “EMS dispatch”, “Remote monitoring”, “Ambulance demand”, “Treatment decision support”, “AED/Station positioning”, and “Research aid” (e.g., patient-trial matching). For each study, we recorded if it used one or more of the following inputs: “ECG”, “Audio/Voice recording”, “EHR (electronic health record) data” (categorical or continuous data, e.g., patient age and sex, presence or absence of symptoms, laboratory tests), “EHR free text”, “Public/Government data” (including weather and population data), “Geospatial/GPS data” (e.g., GPS coordinates), “Time-based data” (e.g. season or month of the year), “Still images” (e.g. X-rays, photos), “Moving images” (e.g. videos of echocardiograms), “Vital signs data” (e.g., blood pressure, heart rate), “Others”. We also noted if multiple input types were used. We analysed application-input pairs by aggregating the total number of studies for each pair and identified any implementation gaps using the evidence map. Given the heterogenous nature of PEC data, we wanted to analyse the trends in multimodal input utilisation and how different inputs are being used in each unique AI application.

## Results

Figure 1 shows the PRISMA flowchart of paper selection. The initial search of the six databases returned 4349 papers and we identified one additional paper through hand searching of included articles. After removing 4072 papers on title and abstract screening, we identified 278 studies for full-text screening, of which 106 studies were included for data extraction and subsequent analysis (15-120).

**Figure 1:**
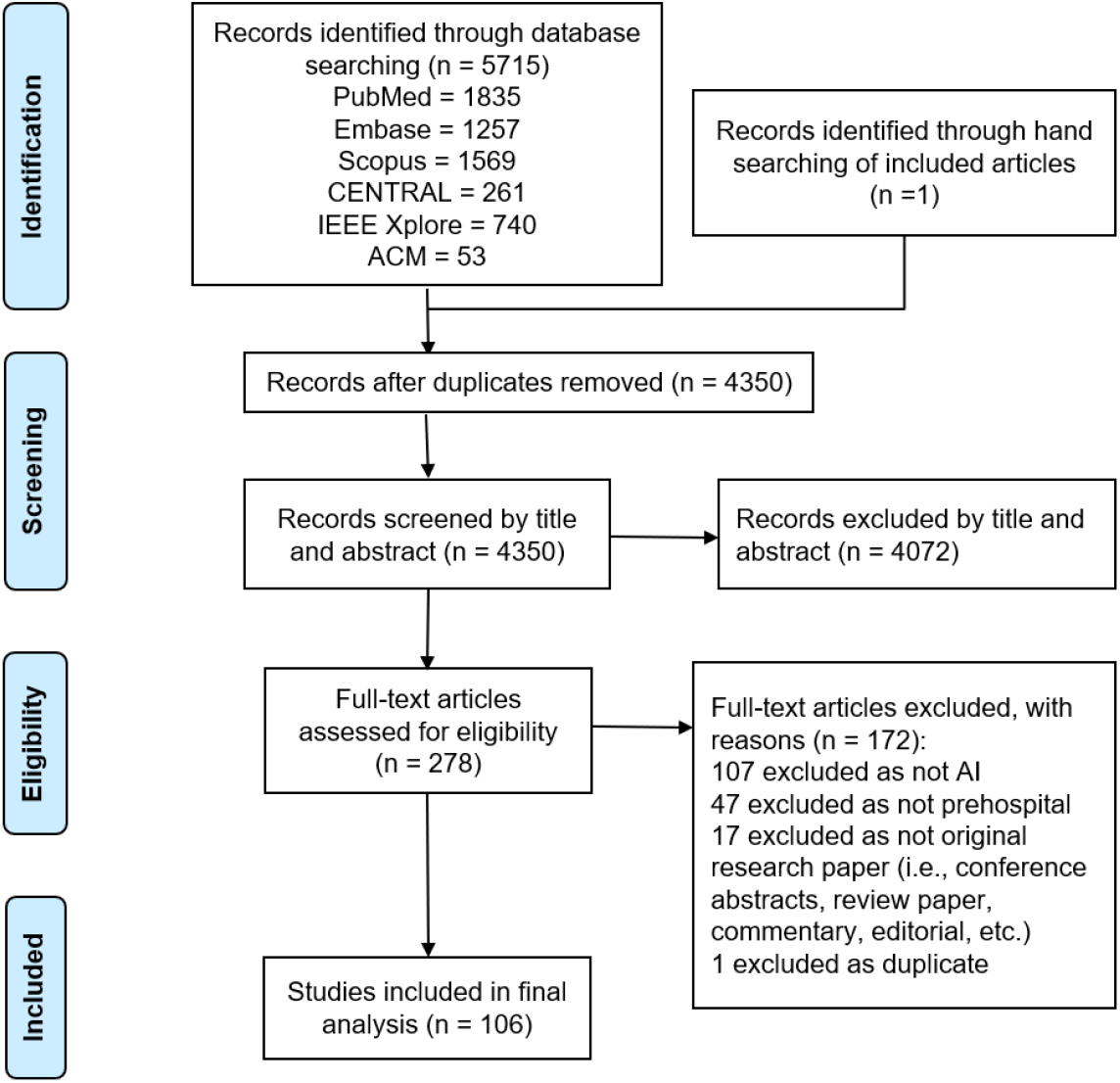
PRISMA flowchart

Table 1 shows the characteristics and methodology of the included studies (for results of individual studies, refer to Supplementary File 3). Datasets from included studies were collected from 25 different countries. Most studies utilised datasets from North America or Europe, with data from United States being the most common (n=46, 32·4%), followed by Sweden (n=12, 11·3%), Norway (n=11, 10·4%), Japan (n=9, 8·5%), and the United Kingdom (n=8, 7·5%).

**Table 1:** Study characteristics and methodology

| Variable | n (%) |
| --- | --- |
| **AI Type** |  |
| Classification tree | 2 (1·9) |
| Decision tree | 6 (5·7) |
| Gradient boosted algorithm | 2 (1·9) |
| Linear classifier | 1 (0·9) |
| Mixed models | 40 (37·7) |
| NLP | 1 (0·9) |
| Neural network | 28 (26·4) |
| Support vector machine | 6 (5·7) |
| Random forest | 14 (13·2) |
| Combined | 10 (9·4) |
| Comparison | 30 (28·3) |
| Other | 2 (1·9) |
| **Comparator** |  |
| Clinical decision tools | 10 (9·4) |
| Human comparator | 5 (4·7) |
| Other AI | 22 (20·8) |
| Non-AI statistical models | 10 (9·4) |
| Other | 2 (1·9) |
| None | 57 (53·8) |
| **Study Type** |  |
| Development | 4 (3·8) |
| Development + validation | 93 (87·7) |
| Predictor finding | 4 (3·8) |
| Validation only | 5 (4·7) |
| **Study Design** |  |
| Retrospective cohort | 88 (83·0) |
| Prospective cohort | 17 (16·0) |
| Randomised controlled trial | 1 (0·9) |
| **Calibration** |  |
| Not reported | 96 (90·6) |
| Reported | 10 (9·4) |
| **Country (of dataset origin)** |  |
| USA | 46 (43·4) |
| Sweden | 12 (11·3) |
| Netherlands | 3 (2·8) |
| Austria | 4 (3·8) |
| France | 5 (4·7) |
| Norway | 11 (10·4) |
| UK | 8 (7·5) |
| Denmark | 2 (1·9) |
| Canada | 1 (0·9) |
| China | 4 (3·8) |
| Czech Republic | 2 (1·9) |
| Finland | 2 (1·9) |
| Japan | 9 (8·5) |
| Greece | 2 (1·9) |
| Italy | 3 (2·8) |
| Ireland | 1 (0·9) |
| Korea | 2 (1·9) |
| Hong Kong | 1 (0·9) |
| Germany | 1 (0·9) |
| Mexico | 1 (0·9) |
| South Africa | 3 (2·8) |
| Taiwan | 3 (2·8) |
| Spain | 2 (1·9) |
| Slovenia | 1 (0·9) |
| Singapore | 1 (0·9) |
| **Inputs** |  |
| ECG | 40 (37·7) |
| Audio | 5 (4·7) |
| EHR | 53 (50·0) |
| Text | 8 (7·5) |
| Public | 5 (4·7) |
| Temporal | 5 (4·7) |
| GIS | 8 (7·5) |
| Image | 1 (0·9) |
| Video | 0 (0·0) |
| Vitals | 15 (14·2) |
| Multiple | 37 (34·9) |
| Other | 7 (6·6) |

The majority of included studies utilised a retrospective cohort (n=88, 83·0%), with a few prospective cohorts (n=17, 16·0%). Only one (0·9%) study was evaluated using a randomised controlled trial.

Figure 2 shows the frequency of each TRIPOD type, with explanations of each type. Most studies were internally validated (n=96, 90·6%). The most common TRIPOD classification was 1B (n=45, 42·5%), where validation was done using re-sampling techniques. Type 2A (n=27, 25·5%) and 2B (n=20, 18·9%) were the next most common. Only 3·8% of studies (n=4) were type 1A and did not perform validation. External validation is more robust but only 8·5% (n=9) of studies used it; 3·8% (n=4) were type 3, models were developed and validated on separate data, and 4·7% (n=5) were type 4, where existing models were evaluated on separate data. One study (71) was not classifiable as it was a predictor finding study that did not create a predictive model. Calibration was evaluated in only 9·4% (n=10) of studies.

**Figure 2:**
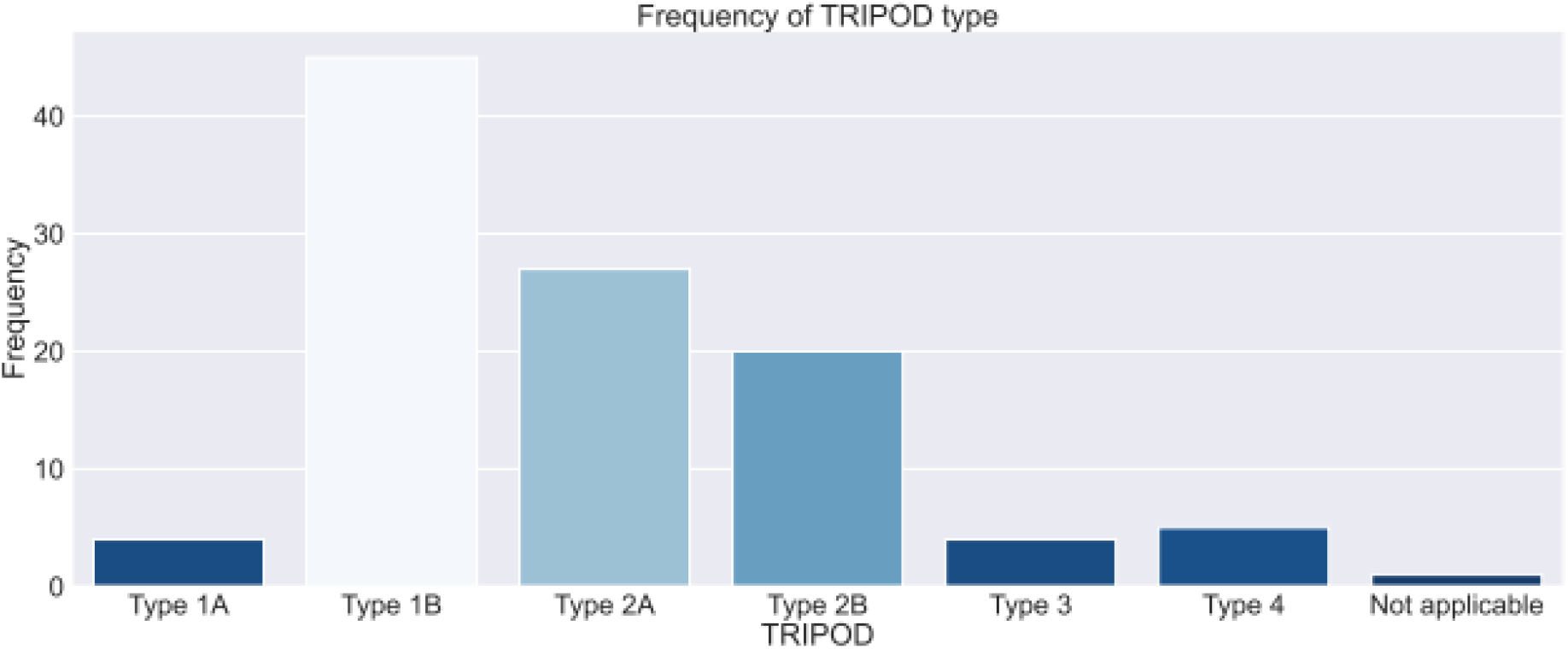
Frequency of TRIPOD type

Included studies used a variety of AI types, with 37·7% of studies (n=40) using multiple models. Of these, 10 studies (9·3%) combined models and 30 (28·3%) developed and compared multiple models. For studies with a single AI model, 28 (26·4%) used neural networks, 14 (14·2%) used random forest, six (5·7%) used decision trees, six (5·7%) used support vector machines, two (1·9%) used classification trees, two (1·9%) used gradient boosted algorithms, one (0·9%) utilised a linear classifier, and one (0·9%) employed natural language processing.

Figure 3 shows the number of studies published per year, stratified according to AI application. Triage/Prognostication (n=52, 49·1%) represented the majority of applications from 2015 onwards, with 57·6% in 2021. CPR/AED optimisation publications (n=26, 24·5%) also increased significantly from 2016, with 38·1% in 2020. The number of publications on AI in PEC increased sharply in 2019, peaking at 33 in 2021, compared to one in 2015. From 2017 to 2021, the diversity of AI applications also increased from two to six out of nine application types. Notably, remote monitoring (n=2, 1·9%), research aid (n=1, 0·9%), AED/station positioning (n=1, 0·9%) and treatment decision support (n=2, 1·9%) were underrepresented in the included studies.

**Figure 3:**
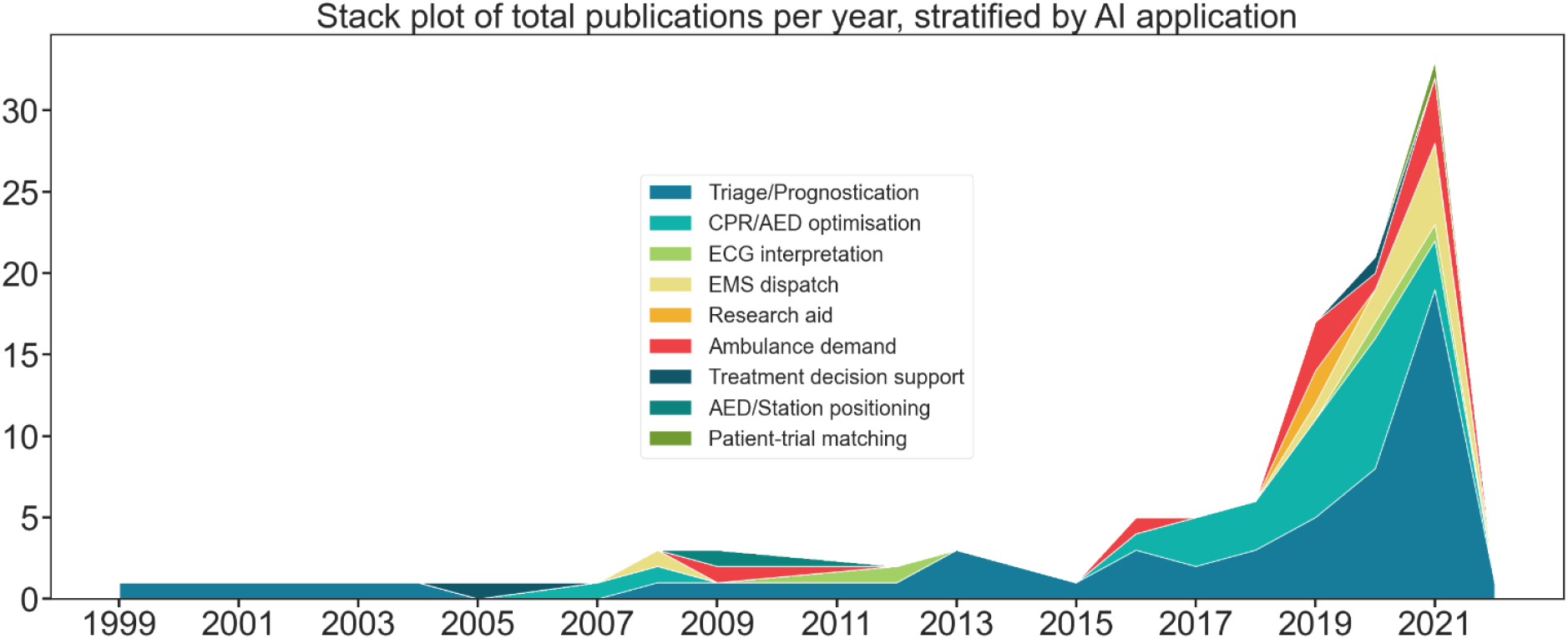
Stack plot of total publications per year, stratified by AI application

Figure 4 shows the performance of AI models against comparators in included studies. In this review, we defined a comparator as any benchmark of performance for the best-performing AI model in the study. AI and non-AI models developed as part of the same study were excluded as comparators. Fifty-seven (53·8%) studies did not use a comparator, 22 (20·8%) used other previously developed AI models, 10 (9·4%) used existing clinical decision tools, ten (9·4%) used non-AI statistical models, and five (4·7%) used human comparators. Two studies (1·9%) used comparators not included in these categories, such as baseline polices and baseline decision rules (31, 67).

**Figure 4:**
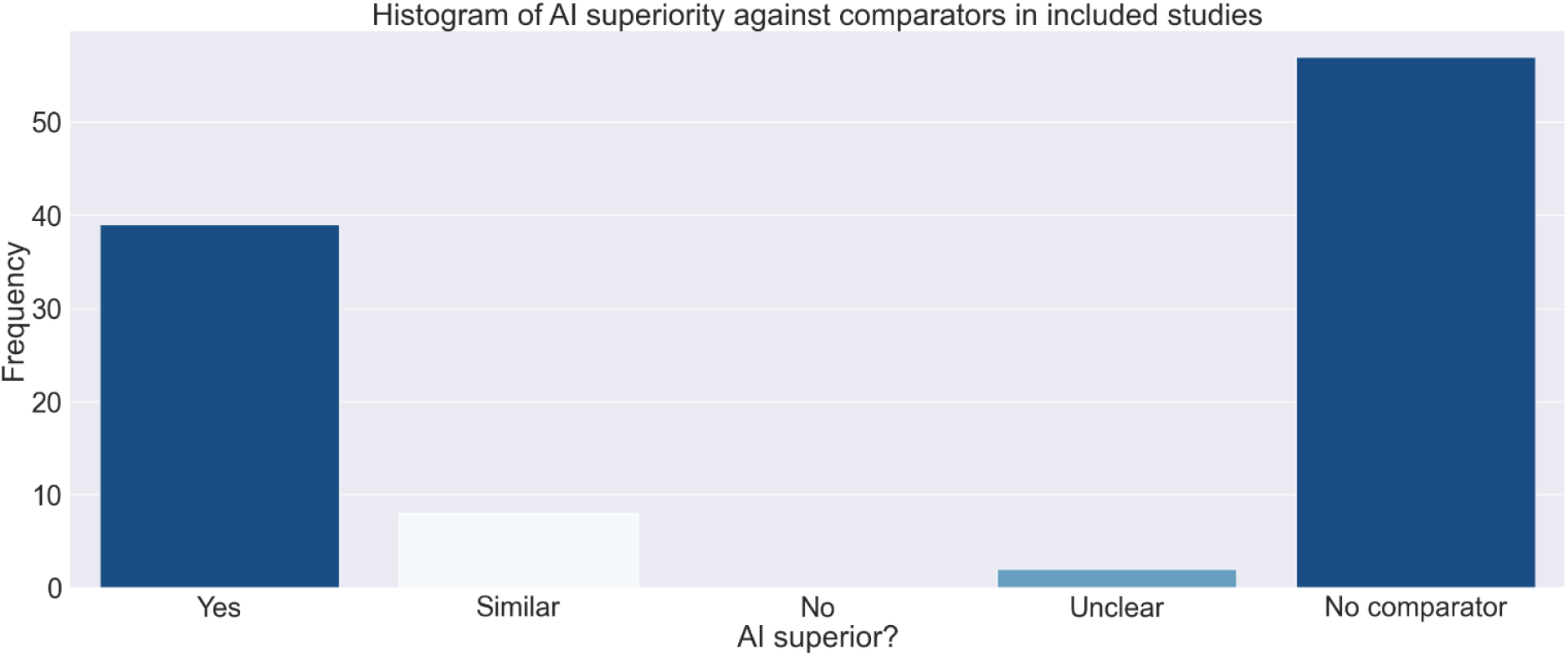
Histogram of AI superiority against comparators in included studies

**Figure 5:**
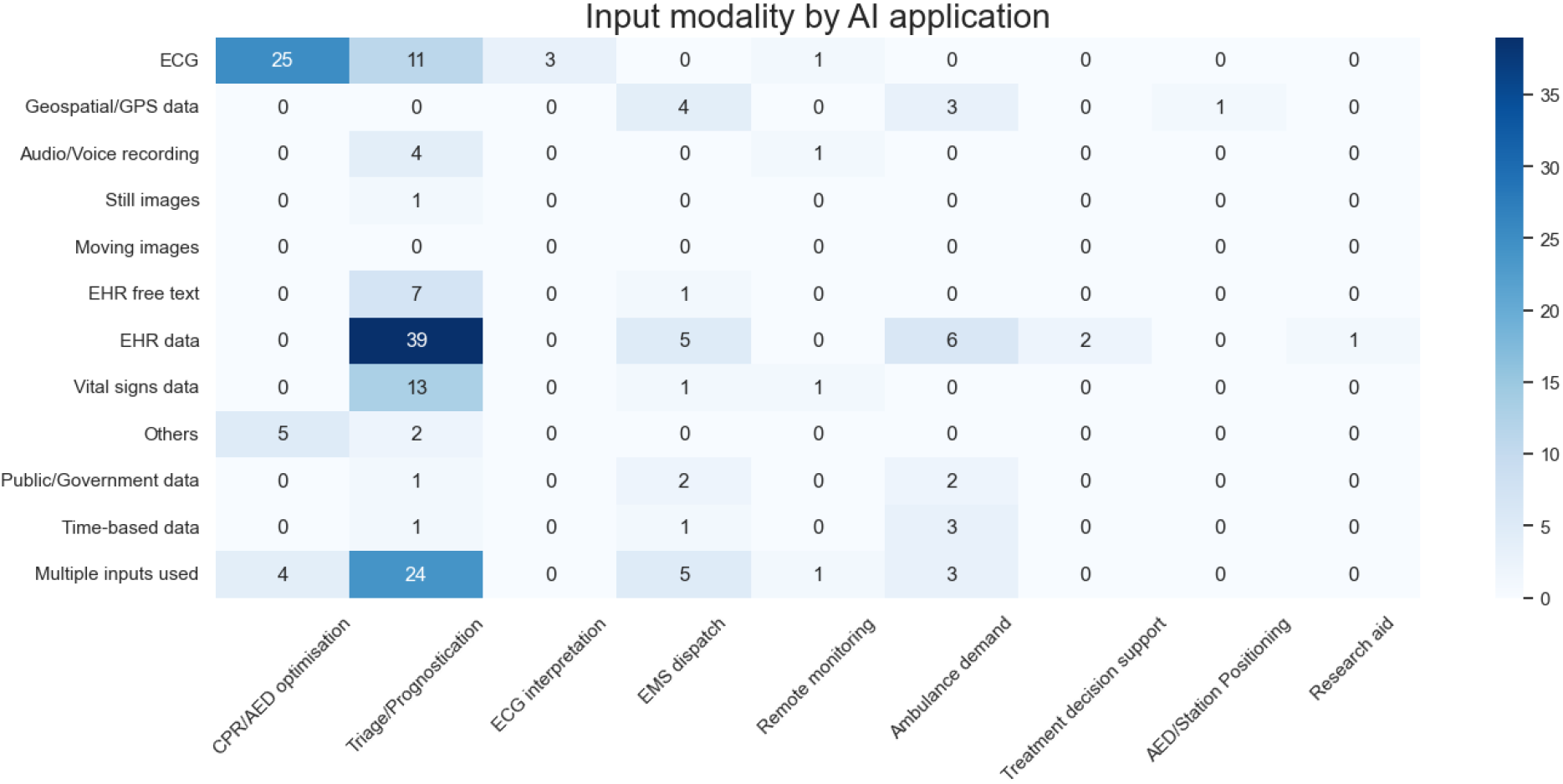
Evidence map of input modality by AI application

Among 49 studies that used comparator against AI, AI was superior in 39 (79·6%) and not statistically different in 8 (16·3%). Results were unclear in two (4·1%) studies. No AI model reported worse performance than the comparator.

We performed evidence map analysis to visualise the landscape of prehospital AI research and identify gaps, as has been demonstrated in previous reviews for AI in COVID-19 research (12). Figure 4 shows the evidence map of input modality compared against application type. CPR/AED optimisation relies heavily on ECG (25 out of 26) as an input and tends to be single input (22 out of 26). Triage/Prognostication leaned more towards having multiple inputs (24 out of 52), with the majority (39 out of 52) using EHR. Inputs such as ECG (40 out of 106), EHR (53 out of 106) and vitals (15 out of 106) were among the most used. The minority of studies utilised multimodal inputs (n=37, 34·9%), with few models using text (n=8), audio (n=5), images (n=1), or videos (n=0) as inputs. Seven studies used inputs that did not fall into one of our predefined categories; these inputs included capnography (38, 107), thoracic impedance (19, 38, 100, 101, 117), and accelerometer-based chest compression depth data (98).

## Discussion

Recently, interest in AI and its applications in PEC has been rapidly growing, with diverse applications promising improvements to PEC systems globally. In this scoping review, we present the first overview of AI applications in PEC settings, including an evidence map analysis of current implementation gaps. AI applications in PEC have been reported to be superior to clinicians or non-AI algorithms, particularly in predictive tasks. Applications of AI in PEC are also diverse, including triage, resource optimisation for dispatch, and geospatial optimisation for stations and AEDs. However, gaps remain in the utilisation of multimodal inputs and novel input modalities such as text, audio, images, and video. In this discussion, we summarise the main findings of our review and provide insight into the potential benefits and challenges of AI in prehospital care.

We found that, like other areas of medicine, the most prevalent application of AI in PEC is triage and prognostication, in the form of diagnostic and prognostic predictive models. These models have the potential to excel as rapid, objective tools for triage and prognostication in PEC settings, where clinician decision making is often time sensitive. Prognostic models help identify patients who may be at high risk for poor outcomes, allowing for earlier intervention and management. Works by Liu et al. (80, 81) demonstrate how the combination of different features such as vital signs and heart rate variability and complexity in a ML prognostic model can provide an accurate estimation of risk in the prehospital setting. These models based on neural networks and multilayer perceptrons can accurately assess the need for lifesaving interventions in trauma patients in real-time. The works of Liu et al. highlight the capability of AI to harness advances in technology and healthcare big data for real-time, continuous monitoring and processing of in-ambulance data, such as vital signs and ECG signals. Similarly, Czap et al. (33) have taken advantage of developments in Mobile Stroke Units (MSUs) and validated an AI algorithm for the prehospital identification of large vessel occlusion using MSU CT angiograms.

Another major domain in prehospital prognostication is out-of-hospital cardiac arrest. AI algorithms have been employed in the prediction of defibrillation success, as well as short- and long-term outcomes following OHCA. Patient outcomes may be improved with further research on the utility of these models in influencing early intervention and other treatment decisions in certain high-risk patients after OHCA.

AI has also been used in various optimisation problems within PEC settings. Several studies have demonstrated the feasibility of AI-assisted dispatch systems to significantly improve response times and increase the efficiency of EMS operations. These studies mainly employ AI for the prediction of travel time (21, 27, 31, 36, 92, 115) and ambulance demand (29, 48, 49, 57, 69, 78, 79, 85, 99, 106), which can assist with the generation of spatial coverage plans for EMS stations (36). Similarly, Mackle et al. (83) used a genetic algorithm to simulate and optimise aerial AED drone positioning for quick access to patients in OHCAs, which may improve long-term outcomes and survival rates.

We found several emerging, novel use cases of AI in PEC. Firstly, Stemerman et al. (112) used clinical notes derived from the EMS to train ML algorithms for patient trial matching, potentially reducing the workload of research nurses and expediting research processes. Also of note is the emerging use of wearable IoT devices. Majumder et al (84) introduced a novel application of AI in pre-hospital patients using a wearable IoT device which signals the users OHCA risk with an approximate accuracy of 95%. Chan et al. (28) investigated contactless detection of cardiac arrest through the integration of AI models that perform real-time classification of agonal breathing into smart IoT devices. With wearable IoT devices becoming more common, model inputs such as ECGs, vital signs, and potentially EHR will also become more readily accessible. With these rich information sources, there is significant potential for applying advanced AI and ML (121, 122) and novel physiological measures (123) for remote continuous monitoring. However, such IoT systems are nascent and require further validation in larger datasets and real-world contexts.

The reported performance of AI applications has been encouraging, with several predictive models achieving areas under the receiver operating characteristic curve (AUROC) greater than 0·9 in their intended discriminatory tasks. However, we caution that these statistics may be optimistic. Many studies were internally validated (TRIPOD type 1A, 1B and 2A) while few were validated by appropriately splitting data temporally or spatially (type 2B) or validated on external datasets from other studies (type 3). Reporting of performance metrics such as calibration was also poor. It is thus uncertain whether the superior discrimination metrics reported in AI studies will translate to efficacy in real-world clinical scenarios which are more dynamic and heterogeneous. Regardless of performance, these AI applications are often the first decision support tools of their kind, with no previous benchmarks or comparators available. These applications represent new opportunities for decision support in triage and prognostication, resource optimisation, and monitoring that have not been possible without AI. Rigorous validation and improved reporting will help to optimise these applications for translation into real-world practice. We recommend that future authors consult AI-specific guidelines such as SPIRIT-AI, CONSORT-AI, and more recently, DECIDE-AI, to guide model development and reporting of results (124, 125).

AI has several advantages over traditional methods in PEC settings. It can effectively analyse and interpret high-dimensional data, such as EHR data, images, and ECG signals (18, 24, 45). AI can also integrate multimodal data (126) and model nonlinear relationships. Shandilya et al. (107) demonstrate this with nonlinear feature extraction and fusion of multimodal capnographic and ECG signal data, resulting in a prediction of defibrillation outcomes with an AUROC of 93·8%. Pirneskoski et al. (95) and Spangler et al. (111)’s AI models for risk prediction of various short-term outcomes outperformed the National Early Warning Scores (NEWS) even when using the same variables, suggesting superior discrimination with nonlinear modelling. Performance was further improved when multimodal data was included (95). Several studies used NLP to analyse multimodal EHR free-text data and speech audio samples for OHCA identification (22, 23, 25) or general triage (42), a task not possible with traditional methods. Nonetheless, the inclusion of multimodal data does not always improve performance (102). Additional data modalities also introduce implementation challenges, such as privacy concerns and data acquisition (126). Currently, multimodal AI is feasible on a small scale, but these challenges and technical limitations prevent the integration of large and diverse data. PEC data is highly multimodal, including ECG signals, ultrasound (127) and CT imaging (128), and image, video, and audio from body worn cameras (129) or wearables (130, 131). With progress in multimodal AI, we anticipate improved performance and greater diversity in PEC AI applications.

Despite clear advantages of AI in predictive performance and versatility, the lack of interpretability is a major barrier to implementation (132). Healthcare professionals are hesitant to accept predictions from AI models without rationale, particularly in high acuity PEC settings. Opaque AI models whose predictions cannot be easily understood, known as ‘black boxes’, raise ethical concerns as they can lead to biased decision making and lack of accountability for any adverse outcomes (133). Thus, researchers may instead opt to use interpretable non-AI methods, such as logistic regression, or less complex AI models (134). An example is Goto et al.’s (46) work with simple, interpretable decision-trees for EMS triage. This solution often, but not always (26, 77, 96, 102, 118), results in poorer discrimination compared to more complex methods like neural networks and deep learning (42, 62, 75). The challenge, then, is appropriately applying AI or non-AI methods in consideration of the clinical context and acceptable limits for performance and interpretability.

A promising solution to model opacity is explainable AI, an approach that seeks to increase AI transparency without compromising performance (135). Explainable AI techniques, such as feature attribution and model agnostic methods, can help practitioners understand the model’s decision-making process and identify potential biases. The shift towards explainable AI enables applications to evolve beyond mere black boxes and serve as valuable decision support tools for practitioners. Yet, at present, not all AI algorithms have suitable explainability methods. In such cases, Ghassemi et al. (136) argue that rigorous validation processes can instil sufficient trust and minimise bias in AI models. While validation processes may serve as a stopgap measure, the field of explainable AI remains a critical area of research for the continued progress and flourishing of AI in PEC settings.

### Limitations

Our study has several limitations. Firstly, we excluded articles on military and disaster medicine, which some may consider relevant to PEC. Our search criteria were also limited to a pre-specified list of AI models which provided clarity to but may have excluded novel forms of AI. Additionally, we only searched for peer-reviewed English language articles, which may have missed grey literature and non-peer-reviewed articles such as conference abstracts. These limitations may have resulted in underrepresentation of AI applications in non-English speaking countries. Indeed, included studies were predominantly from Europe or North America. Given the scoping nature of the review, we also did not conduct a formal risk of bias analysis. However, despite these limitations, our review provides a systematic overview of the current literature on AI applications in PEC.

## Conclusions

AI in PEC is a growing field, with numerous promising applications such as prognostication, demand prediction, resource optimisation, and IoT continuous monitoring systems. While the potential for AI in PEC is promising, it is important to select appropriate use cases for AI applications and not to over-generalise its capabilities. The field of AI in PEC is still in its infancy and more prospective, externally validated studies are needed before AI can progress beyond the proof-of-concept stage to real-world clinical settings.

## Data Availability

All data produced in the present study are available upon reasonable request to the authors

## Contributors

All authors had full access to all the data in the study and had final responsibility for the decision to submit for publication.

Conceptualization: NL; Study design: MLC1, NL; Literature search: MLC1, KM, KT; Data extraction: MLC1, MLC2, HH; Data verification: MLC1, MLC2, HH, KM, KT, HW, MF, FJS, AFWH, MEHO, NL; Formal analysis: MLC1, MLC2, HH; Investigation: MLC1, MLC2, HH, KM, KT, HW, MF, FJS, AFWH, MEHO, NL; Writing—original draft: MLC1, MLC2, HH; Writing—review and editing: MLC1, MLC2, HH, KM, KT, HW, MF, FJS, AFWH, MEHO, NL.

## Data sharing statement

All data collected for this systematic review, including search strategy and data extraction sheets, are available immediately after publication and are either published as supplementary material or can be accessed through the corresponding author.

## References

1. Jiang F, Jiang Y, Zhi H, Dong Y, Li H, Ma S, et al. Artificial intelligence in healthcare: past, present and future. Stroke and vascular neurology. 2017;2(4):230–43.

2. Yu K-H, Beam AL, Kohane IS. Artificial intelligence in healthcare. Nature Biomedical Engineering. 2018;2(10):719–31.

3. Bizopoulos P, Koutsouris D. Deep Learning in Cardiology. IEEE Reviews in Biomedical Engineering. 2019;12:168–93.

4. Ting DSW, Pasquale LR, Peng L, Campbell JP, Lee AY, Raman R, et al. Artificial intelligence and deep learning in ophthalmology. British Journal of Ophthalmology. 2019;103(2):167–75.

5. Liu N, Zhang Z, Ho AFW, Ong MEH. Artificial intelligence in emergency medicine. Journal of Emergency and Critical Care Medicine. 2018;2:82.

6. Kirubarajan A, Taher A, Khan S, Masood S. Artificial intelligence in emergency medicine: A scoping review. Journal of the American College of Emergency Physicians Open. 2020;n/a(n/a).

7. Habibzadeh H, Dinesh K, Shishvan OR, Boggio-Dandry A, Sharma G, Soyata T. A Survey of Healthcare Internet of Things (HIoT): A Clinical Perspective. IEEE Internet of Things Journal. 2020;7(1):53–71.

8. Stewart J, Sprivulis P, Dwivedi G. Artificial intelligence and machine learning in emergency medicine. Emergency medicine Australasia : EMA. 2018.

9. Berlyand Y, Raja AS, Dorner SC, Prabhakar AM, Sonis JD, Gottumukkala RV, et al. How artificial intelligence could transform emergency department operations. The American journal of emergency medicine. 2018;36(8):1515–7.

10. Fernandes M, Vieira SM, Leite F, Palos C, Finkelstein S, Sousa JMC. Clinical Decision Support Systems for Triage in the Emergency Department using Intelligent Systems: a Review. Artificial Intelligence in Medicine. 2020;102:101762.

11. Chee ML, Ong MEH, Siddiqui FJ, Zhang Z, Lim SL, Ho AFW, et al. Artificial Intelligence Applications for COVID-19 in Intensive Care and Emergency Settings: A Systematic Review. Int J Environ Res Public Health. 2021;18(9).

12. Liu N, Chee ML, Niu C, Pek PP, Siddiqui FJ, Ansah JP, et al. Coronavirus disease 2019 (COVID-19): an evidence map of medical literature. BMC Medical Research Methodology. 2020;20(1):177.

13. Saran A, White H. Evidence and gap maps: a comparison of different approaches. Campbell Systematic Reviews. 2018;14(1):1–38.

14. Collins GS, Reitsma JB, Altman DG, Moons KGM. Transparent Reporting of a Multivariable Prediction Model for Individual Prognosis or Diagnosis (TRIPOD). Circulation. 2015;131(2):211–9.

15. Acharya UR, Fujita H, Oh SL, Raghavendra U, Tan JH, Adam M, et al. Automated identification of shockable and non-shockable life-threatening ventricular arrhythmias using convolutional neural network. Future Generation Computer Systems. 2018;79:952–9.

16. Ajumobi O, Verdugo SR, Labus B, Reuther P, Lee B, Koch B, et al. Identification of Non-Fatal Opioid Overdose Cases Using 9-1-1 Computer Assisted Dispatch and Prehospital Patient Clinical Record Variables. Prehospital Emergency Care. 2021;26(6):818–28.

17. Al-Dury N, Ravn-Fischer A, Hollenberg J, Israelsson J, Nordberg P, Strömsöe A, et al. Identifying the relative importance of predictors of survival in out of hospital cardiac arrest: a machine learning study. Scandinavian Journal of Trauma, Resuscitation and Emergency Medicine. 2020;28(1).

18. Al-Zaiti S, Besomi L, Bouzid Z, Faramand Z, Frisch S, Martin-Gill C, et al. Machine learning-based prediction of acute coronary syndrome using only the pre-hospital 12-lead electrocardiogram. Nat Commun. 2020;11(1):3966.

19. Alonso E, Irusta U, Aramendi E, Daya MR. A Machine Learning Framework for Pulse Detection During Out-of-Hospital Cardiac Arrest. IEEE Access. 2020;8:161031–41.

20. Anthony T, Mishra AK, Stassen W, Son J. The Feasibility of Using Machine Learning to Classify Calls to South African Emergency Dispatch Centres According to Prehospital Diagnosis, by Utilising Caller Descriptions of the Incident. Healthcare. 2021;9(9).

21. Arcolezi HH, Cerna S, Guyeux C, Couchot J-F. Preserving Geo-Indistinguishability of the Emergency Scene to Predict Ambulance Response Time. Mathematical and Computational Applications. 2021;26(3):56.

22. Blomberg SN, Christensen HC, Lippert F, Ersboll AK, Torp-Petersen C, Sayre MR, et al. Effect of Machine Learning on Dispatcher Recognition of Out-of-Hospital Cardiac Arrest During Calls to Emergency Medical Services: A Randomized Clinical Trial. JAMA Netw Open. 2021;4(1):e2032320.

23. Blomberg SN, Folke F, Ersboll AK, Christensen HC, Torp-Pedersen C, Sayre MR, et al. Machine learning as a supportive tool to recognize cardiac arrest in emergency calls. Resuscitation. 2019;138:322–9.

24. Bouzid Z, Faramand Z, Gregg RE, Helman S, Martin-Gill C, Saba S, et al. Novel ECG features and machine learning to optimize culprit lesion detection in patients with suspected acute coronary syndrome. J Electrocardiol. 2021;69S:31–7.

25. Byrsell F, Claesson A, Ringh M, Svensson L, Jonsson M, Nordberg P, et al. Machine learning can support dispatchers to better and faster recognize out-of-hospital cardiac arrest during emergency calls: A retrospective study. Resuscitation. 2021;162:218–26.

26. Candefjord S, Sheikh Muhammad A, Bangalore P, Buendia R. On Scene Injury Severity Prediction (OSISP) machine learning algorithms for motor vehicle crash occupants in US. Journal of Transport & Health. 2021;22:101124.

27. Cerna S, Arcolezi HH, Guyeux C, Royer-Fey G, Chevallier C. Machine learning-based forecasting of firemen ambulances’ turnaround time in hospitals, considering the COVID-19 impact. Appl Soft Comput. 2021;109:107561.

28. Chan J, Rea T, Gollakota S, Sunshine JE. Contactless cardiac arrest detection using smart devices. npj Digital Medicine. 2019;2(1):52.

29. Chen AY, Lu TY, Ma MH, Sun WZ. Demand Forecast Using Data Analytics for the Preallocation of Ambulances. IEEE J Biomed Health Inform. 2016;20(4):1178–87.

30. Chen Z, Zhang R, Xu F, Gong X, Shi F, Zhang M, et al. Novel Prehospital Prediction Model of Large Vessel Occlusion Using Artificial Neural Network. Frontiers in Aging Neuroscience. 2018;10.

31. Chu J, Leung KHB, Snobelen P, Nevils G, Drennan IR, Cheskes S, et al. Machine learning-based dispatch of drone-delivered defibrillators for out-of-hospital cardiac arrest. Resuscitation. 2021;162:120–7.

32. Coult J, Rea TD, Blackwood J, Kudenchuk PJ, Liu C, Kwok H. A method to predict ventricular fibrillation shock outcome during chest compressions. Computers in Biology and Medicine. 2021;129.

33. Czap AL, Bahr-Hosseini M, Singh N, Yamal JM, Nour M, Parker S, et al. Machine Learning Automated Detection of Large Vessel Occlusion From Mobile Stroke Unit Computed Tomography Angiography. Stroke. 2022;53(5):1651–6.

34. Davis DP, Peay J, Good B, Sise MJ, Kennedy F, Eastman AB, et al. Air Medical Response to Traumatic Brain Injury: A Computer Learning Algorithm Analysis. Journal of Trauma: Injury, Infection & Critical Care. 2008;64(4):889–97.

35. Davis DP, Peay J, Sise MJ, Vilke GM, Kennedy F, Eastman AB, et al. The Impact of Prehospital Endotracheal Intubation on Outcome in Moderate to Severe Traumatic Brain Injury. The Journal of Trauma: Injury, Infection, and Critical Care. 2005;58(5):933–9.

36. Dolejš M, Purchard J, Javorčák A. Generating a spatial coverage plan for the emergency medical service on a regional scale: Empirical versus random forest modelling approach. Journal of Transport Geography. 2020;89:102889.

37. Duceau B, Alsac JM, Bellenfant F, Mailloux A, Champigneulle B, Favé G, et al. Prehospital triage of acute aortic syndrome using a machine learning algorithm. British Journal of Surgery. 2020;107(8):995–1003.

38. Elola A, Aramendi E, Irusta U, Berve PO, Wik L. Multimodal Algorithms for the Classification of Circulation States During Out-of-Hospital Cardiac Arrest. IEEE Transactions on Biomedical Engineering. 2021;68(6):1913–22.

39. Elola A, Aramendi E, Irusta U, Del Ser J, Alonso E, Daya M. ECG-based pulse detection during cardiac arrest using random forest classifier. Medical & Biological Engineering & Computing. 2018;57(2):453–62.

40. Elola A, Aramendi E, Irusta U, Picón A, Alonso E, Owens P, et al. Deep Neural Networks for ECG-Based Pulse Detection during Out-of-Hospital Cardiac Arrest. Entropy. 2019;21(3).

41. Elola A, Aramendi E, Rueda E, Irusta U, Wang H, Idris A. Towards the Prediction of Rearrest during Out-of-Hospital Cardiac Arrest. Entropy. 2020;22(7).

42. Ferri P, Saez C, Felix-De Castro A, Juan-Albarracin J, Blanes-Selva V, Sanchez-Cuesta P, et al. Deep ensemble multitask classification of emergency medical call incidents combining multimodal data improves emergency medical dispatch. Artif Intell Med. 2021;117:102088.

43. Figuera C, Irusta U, Morgado E, Aramendi E, Ayala U, Wik L, et al. Machine Learning Techniques for the Detection of Shockable Rhythms in Automated External Defibrillators. PLoS One. 2016;11(7):e0159654.

44. Follin A, Jacqmin S, Chhor V, Bellenfant F, Robin S, Guinvarc’h A, et al. Tree-based algorithm for prehospital triage of polytrauma patients. Injury. 2016;47(7):1555–61.

45. Forberg JL, Khoshnood A, Green M, Ohlsson M, Bjork J, Jovinge S, et al. An artificial neural network to safely reduce the number of ambulance ECGs transmitted for physician assessment in a system with prehospital detection of ST elevation myocardial infarction. Scand J Trauma Resusc Emerg Med. 2012;20:8.

46. Goto Y, Maeda T, Goto Y. Decision-tree model for predicting outcomes after out-of-hospital cardiac arrest in the emergency department. Critical Care. 2013;17(4):R133.

47. Goto Y, Maeda T, Nakatsu-Goto Y. Decision tree model for predicting long-term outcomes in children with out-of-hospital cardiac arrest: a nationwide, population-based observational study. Critical Care. 2014;18(3).

48. Grekousis G, Liu Y. Where will the next emergency event occur? Predicting ambulance demand in emergency medical services using artificial intelligence. Computers, Environment and Urban Systems. 2019;76:110–22.

49. Grekousis G, Photis YN. Analyzing High-Risk Emergency Areas with GIS and Neural Networks: The Case of Athens, Greece. The Professional Geographer. 2014;66(1):124–37.

50. Hajeb-M S, Cascella A, Valentine M, Chon KH. Deep Neural Network Approach for Continuous ECG-Based Automated External Defibrillator Shock Advisory System During Cardiopulmonary Resuscitation. Journal of the American Heart Association. 2021;10(6).

51. Harford S, Darabi H, Del Rios M, Majumdar S, Karim F, Vanden Hoek T, et al. A machine learning based model for Out of Hospital cardiac arrest outcome classification and sensitivity analysis. Resuscitation. 2019;138:134–40.

52. Hayashi Y, Shimada T, Hattori N, Shimazui T, Yoshida Y, Miura RE, et al. A prehospital diagnostic algorithm for strokes using machine learning: a prospective observational study. Scientific Reports. 2021;11(1).

53. He M, Gong Y, Li Y, Mauri T, Fumagalli F, Bozzola M, et al. Combining multiple ECG features does not improve prediction of defibrillation outcome compared to single features in a large population of out-of-hospital cardiac arrests. Critical Care. 2015;19(1).

54. He M, Lu Y, Zhang L, Zhang H, Gong Y, Li Y. Combining Amplitude Spectrum Area with Previous Shock Information Using Neural Networks Improves Prediction Performance of Defibrillation Outcome for Subsequent Shocks in Out-Of-Hospital Cardiac Arrest Patients. PLoS One. 2016;11(2):e0149115.

55. Hirano Y, Kondo Y, Sueyoshi K, Okamoto K, Tanaka H. Early outcome prediction for out-of-hospital cardiac arrest with initial shockable rhythm using machine learning models. Resuscitation. 2021;158:49–56.

56. Hotradat M, Balasundaram K, Masse S, Nair K, Nanthakumar K, Umapathy K. Empirical mode decomposition based ECG features in classifying and tracking ventricular arrhythmias. Computers in Biology and Medicine. 2019;112.

57. Huang H, Jiang M, Ding Z, Zhou M. Forecasting Emergency Calls With a Poisson Neural Network-Based Assemble Model. IEEE Access. 2019;7:18061–9.

58. Isasi I, Irusta U, Aramendi E, Eftestøl T, Kramer-Johansen J, Wik L. Rhythm Analysis during Cardiopulmonary Resuscitation Using Convolutional Neural Networks. Entropy. 2020;22(6).

59. Isasi I, Irusta U, Aramendi E, Olsen JA, Wik L. Shock decision algorithm for use during load distributing band cardiopulmonary resuscitation. Resuscitation. 2021;165:93–100.

60. Isasi I, Irusta U, Bahrami Rad A, Aramendi E, Zabihi M, Eftestol T, et al. Automatic Cardiac Rhythm Classification With Concurrent Manual Chest Compressions. IEEE Access. 2019;7:115147–59.

61. Isasi I, Irusta U, Elola A, Aramendi E, Ayala U, Alonso E, et al. A Machine Learning Shock Decision Algorithm for Use During Piston-Driven Chest Compressions. IEEE Transactions on Biomedical Engineering. 2019;66(6):1752–60.

62. Ivanović MD, Hannink J, Ring M, Baronio F, Vukčević V, Hadžievski L, et al. Predicting defibrillation success in out-of-hospital cardiac arrested patients: Moving beyond feature design. Artif Intell Med. 2020;110:101963.

63. Ivanović MD, Ring M, Baronio F, Calza S, Vukčević V, Hadžievski L, et al. ECG derived feature combination versus single feature in predicting defibrillation success in out-of-hospital cardiac arrested patients. Biomedical Physics & Engineering Express. 2018;5(1).

64. Jaureguibeitia X, Irusta U, Aramendi E, Owens PC, Wang HE, Idris AH. Automatic Detection of Ventilations During Mechanical Cardiopulmonary Resuscitation. IEEE Journal of Biomedical and Health Informatics. 2020;24(9):2580–8.

65. Jaureguibeitia X, Zubia G, Irusta U, Aramendi E, Chicote B, Alonso D, et al. Shock Decision Algorithms for Automated External Defibrillators Based on Convolutional Networks. IEEE Access. 2020;8:154746–58.

66. Jekova I, Krasteva V. Optimization of End-to-End Convolutional Neural Networks for Analysis of Out-of-Hospital Cardiac Arrest Rhythms during Cardiopulmonary Resuscitation. Sensors. 2021;21(12).

67. Ji S, Zheng Y, Wang Z, Li T. A Deep Reinforcement Learning-Enabled Dynamic Redeployment System for Mobile Ambulances. Proceedings of the ACM on Interactive, Mobile, Wearable and Ubiquitous Technologies. 2019;3(1):1–20.

68. Jiang Y-J, Ma MH-M, Sun W-Z, Chang K-W, Abbod MF, Shieh J-S. Ensembled neural networks applied to modeling survival rate for the patients with out-of-hospital cardiac arrest. Artificial Life and Robotics. 2012;17(2):241–4.

69. Jin R, Xia T, Liu X, Murata T, Kim KS. Predicting Emergency Medical Service Demand With Bipartite Graph Convolutional Networks. IEEE Access. 2021;9:9903–15.

70. Kang D-Y, Cho K-J, Kwon O, Kwon J-m, Jeon K-H, Park H, et al. Artificial intelligence algorithm to predict the need for critical care in prehospital emergency medical services. Scandinavian Journal of Trauma, Resuscitation and Emergency Medicine. 2020;28(1).

71. Kao J-H, Chan T-C, Lai F, Lin B-C, Sun W-Z, Chang K-W, et al. Spatial analysis and data mining techniques for identifying risk factors of Out-of-Hospital Cardiac Arrest. International Journal of Information Management. 2017;37(1):1528–38.

72. Kim D, You S, So S, Lee J, Yook S, Jang DP, et al. A data-driven artificial intelligence model for remote triage in the prehospital environment. PLoS One. 2018;13(10):e0206006.

73. Krasteva V, Ménétré S, Didon J-P, Jekova I. Fully Convolutional Deep Neural Networks with Optimized Hyperparameters for Detection of Shockable and Non-Shockable Rhythms. Sensors. 2020;20(10).

74. Krizmaric M, Verlic M, Stiglic G, Grmec S, Kokol P. Intelligent analysis in predicting outcome of out-of-hospital cardiac arrest. Computer Methods and Programs in Biomedicine. 2009;95(2):S22–S32.

75. Kwon J-m, Jeon K-H, Kim HM, Kim MJ, Lim S, Kim K-H, et al. Deep-learning-based out-of-hospital cardiac arrest prognostic system to predict clinical outcomes. Resuscitation. 2019;139:84–91.

76. Lammers D, Marenco C, Morte K, Conner J, Williams J, Bax T, et al. Machine Learning for Military Trauma: Novel Massive Transfusion Predictive Models in Combat Zones. Journal of Surgical Research. 2022;270:369–75.

77. Larsson A, Berg J, Gellerfors M, Gerdin Wärnberg M. The advanced machine learner XGBoost did not reduce prehospital trauma mistriage compared with logistic regression: a simulation study. BMC Medical Informatics and Decision Making. 2021;21(1):192.

78. Lee H, Lee T. Demand modelling for emergency medical service system with multiple casualties cases: k-inflated mixture regression model. Flexible Services and Manufacturing Journal. 2021;33(4):1090–115.

79. Lin AX, Ho AFW, Cheong KH, Li Z, Cai W, Chee ML, et al. Leveraging Machine Learning Techniques and Engineering of Multi-Nature Features for National Daily Regional Ambulance Demand Prediction. Int J Environ Res Public Health. 2020;17(11).

80. Liu NT, Holcomb JB, Wade CE, Batchinsky AI, Cancio LC, Darrah MI, et al. Development and validation of a machine learning algorithm and hybrid system to predict the need for life-saving interventions in trauma patients. Med Biol Eng Comput. 2014;52(2):193–203.

81. Liu NT, Holcomb JB, Wade CE, Darrah MI, Salinas J. Utility of vital signs, heart rate variability and complexity, and machine learning for identifying the need for lifesaving interventions in trauma patients. Shock. 2014;42(2):108–14.

82. Lo YH, Siu YCA. Predicting Survived Events in Nontraumatic Out-of-Hospital Cardiac Arrest: A Comparison Study on Machine Learning and Regression Models. The Journal of Emergency Medicine. 2021;61(6):683–94.

83. Mackle C, Bond R, Torney H, McBride R, McLaughlin J, Finlay D, et al. A Data-Driven Simulator for the Strategic Positioning of Aerial Ambulance Drones Reaching Out-of-Hospital Cardiac Arrests: A Genetic Algorithmic Approach. IEEE J Transl Eng Health Med. 2020;8:1900410.

84. Majumder AKMJA, ElSaadany YA, Young R, Ucci DR. An Energy Efficient Wearable Smart IoT System to Predict Cardiac Arrest. Advances in Human-Computer Interaction. 2019;2019:1507465.

85. Martin RJ, Mousavi R, Saydam C. Predicting emergency medical service call demand: A modern spatiotemporal machine learning approach. Operations Research for Health Care. 2021;28:100285.

86. Mayampurath A, Parnianpour Z, Richards CT, Meurer WJ, Lee J, Ankenman B, et al. Improving Prehospital Stroke Diagnosis Using Natural Language Processing of Paramedic Reports. Stroke. 2021;52(8):2676–9.

87. Nederpelt CJ, Mokhtari AK, Alser O, Tsiligkaridis T, Roberts J, Cha M, et al. Development of a field artificial intelligence triage tool: Confidence in the prediction of shock, transfusion, and definitive surgical therapy in patients with truncal gunshot wounds. Journal of Trauma and Acute Care Surgery. 2021;90(6):1054–60.

88. Newgard CD, Lewis RJ, Jolly BT. Use of out-of-hospital variables to predict severity of injury in pediatric patients involved in motor vehicle crashes. Annals of Emergency Medicine. 2002;39(5):481–91.

89. Nguyen MT, Nguyen BV, Kim K. Deep Feature Learning for Sudden Cardiac Arrest Detection in Automated External Defibrillators. Scientific Reports. 2018;8(1).

90. Nguyen MT, Shahzad A, Nguyen BV, Kim K. Diagnosis of shockable rhythms for automated external defibrillators using a reliable support vector machine classifier. Biomedical Signal Processing and Control. 2018;44:258–69.

91. Nguyen MT, Van Nguyen B, Kim K. Shockable Rhythm Diagnosis for Automated External Defibrillators Using a Modified Variational Mode Decomposition Technique. IEEE Transactions on Industrial Informatics. 2017;13(6):3037–46.

92. Olave-Rojas D, Nickel S. Modeling a pre-hospital emergency medical service using hybrid simulation and a machine learning approach. Simulation Modelling Practice and Theory. 2021;109:102302.

93. Park JH, Choi J, Lee S, Shin SD, Song KJ. Use of Time-to-Event Analysis to Develop On-Scene Return of Spontaneous Circulation Prediction for Out-of-Hospital Cardiac Arrest Patients. Annals of Emergency Medicine. 2022;79(2):132–44.

94. Picon A, Irusta U, Alvarez-Gila A, Aramendi E, Alonso-Atienza F, Figuera C, et al. Mixed convolutional and long short-term memory network for the detection of lethal ventricular arrhythmia. PLoS One. 2019;14(5):e0216756.

95. Pirneskoski J, Tamminen J, Kallonen A, Nurmi J, Kuisma M, Olkkola KT, et al. Random forest machine learning method outperforms prehospital National Early Warning Score for predicting one-day mortality: A retrospective study. Resusc Plus. 2020;4:100046.

96. Prieto JT, Scott K, McEwen D, Podewils LJ, Al-Tayyib A, Robinson J, et al. The Detection of Opioid Misuse and Heroin Use From Paramedic Response Documentation: Machine Learning for Improved Surveillance. J Med Internet Res. 2020;22(1):e15645.

97. Rad AB, Eftestol T, Engan K, Irusta U, Kvaloy JT, Kramer-Johansen J, et al. ECG-Based Classification of Resuscitation Cardiac Rhythms for Retrospective Data Analysis. IEEE Transactions on Biomedical Engineering. 2017;64(10):2411–8.

98. Rad AB, Eftestøl T, Irusta U, Kvaløy JT, Wik L, Kramer-Johansen J, et al. An automatic system for the comprehensive retrospective analysis of cardiac rhythms in resuscitation episodes. Resuscitation. 2018;122:6–12.

99. Ramgopal S, Westling T, Siripong N, Salcido DD, Martin-Gill C. Use of a metalearner to predict emergency medical services demand in an urban setting. Comput Methods Programs Biomed. 2021;207:106201.

100. Risdal M, Aase SO, Kramer-Johansen J, Eftestol T. Automatic Identification of Return of Spontaneous Circulation During Cardiopulmonary Resuscitation. IEEE Transactions on Biomedical Engineering. 2008;55(1):60–8.

101. Risdal M, Aase SO, Stavland M, Eftestol T. Impedance-Based Ventilation Detection During Cardiopulmonary Resuscitation. IEEE Transactions on Biomedical Engineering. 2007;54(12):2237–45.

102. Sashidhar D, Kwok H, Coult J, Blackwood J, Kudenchuk PJ, Bhandari S, et al. Machine learning and feature engineering for predicting pulse presence during chest compressions. Royal Society Open Science. 2021;8(11).

103. Scerbo M, Radhakrishnan H, Cotton B, Dua A, Del Junco D, Wade C, et al. Prehospital triage of trauma patients using the Random Forest computer algorithm. Journal of Surgical Research. 2014;187(2):371–6.

104. Scheetz LJ, Zhang J, Kolassa J. Classification tree modeling to identify severe and moderate vehicular injuries in young and middle-aged adults. Artificial Intelligence in Medicine. 2009;45(1):1–10.

105. Seki T, Tamura T, Suzuki M. Outcome prediction of out-of-hospital cardiac arrest with presumed cardiac aetiology using an advanced machine learning technique. Resuscitation. 2019;141:128–35.

106. Setzler H, Saydam C, Park S. EMS call volume predictions: A comparative study. Computers & Operations Research. 2009;36(6):1843–51.

107. Shandilya S, Kurz MC, Ward KR, Najarian K. Integration of Attributes from Non-Linear Characterization of Cardiovascular Time-Series for Prediction of Defibrillation Outcomes. PLoS One. 2016;11(1):e0141313.

108. Shirakawa T, Sonoo T, Ogura K, Fujimori R, Hara K, Goto T, et al. Institution-Specific Machine Learning Models for Prehospital Assessment to Predict Hospital Admission: Prediction Model Development Study. JMIR Medical Informatics. 2020;8(10).

109. Siebelt M, Das D, Van Den Moosdijk A, Warren T, Van Der Putten P, Van Der Weegen W. Machine learning algorithms trained with pre-hospital acquired history-taking data can accurately differentiate diagnoses in patients with hip complaints. Acta Orthopaedica. 2021;92(3):254–7.

110. Sivaraman JJ, Proescholdbell SK, Ezzell D, Shanahan ME. Characterizing Opioid Overdoses Using Emergency Medical Services Data. Public Health Reports. 2021;136(1_suppl):62S–71S.

111. Spangler D, Hermansson T, Smekal D, Blomberg H. A validation of machine learning-based risk scores in the prehospital setting. PLoS One. 2019;14(12):e0226518.

112. Stemerman R, Bunning T, Grover J, Kitzmiller R, Patel MD. Identifying Patient Phenotype Cohorts Using Prehospital Electronic Health Record Data. Prehospital Emergency Care. 2022;26(1):78–88.

113. Tamminen J, Kallonen A, Hoppu S, Kalliomäki J. Machine learning model predicts short-term mortality among prehospital patients: A prospective development study from Finland. Resuscitation Plus. 2021;5.

114. Tignanelli CJ, Silverman GM, Lindemann EA, Trembley AL, Gipson JC, Beilman G, et al. Natural language processing of prehospital emergency medical services trauma records allows for automated characterization of treatment appropriateness. Journal of Trauma and Acute Care Surgery. 2020;88(5):607–14.

115. Torres N, Trujillo L, Maldonado Y, Vera C. Correction of the travel time estimation for ambulances of the red cross Tijuana using machine learning. Comput Biol Med. 2021;137:104798.

116. Uchida K, Kouno J, Yoshimura S, Kinjo N, Sakakibara F, Araki H, et al. Development of Machine Learning Models to Predict Probabilities and Types of Stroke at Prehospital Stage: the Japan Urgent Stroke Triage Score Using Machine Learning (JUST-ML). Translational Stroke Research. 2021;13(3):370–81.

117. Urteaga J, Aramendi E, Elola A, Irusta U, Idris A. A Machine Learning Model for the Prognosis of Pulseless Electrical Activity during Out-of-Hospital Cardiac Arrest. Entropy. 2021;23(7).

118. Yang L, Liu Q, Zhao Q, Zhu X, Wang L. Machine learning is a valid method for predicting prehospital delay after acute ischemic stroke. Brain Behav. 2020;10(10):e01794.

119. Yang Z, Yang Z, Lu W, Harrison RG, Eftestøl T, Steen PA. A probabilistic neural network as the predictive classifier of out-of-hospital defibrillation outcomes. Resuscitation. 2005;64(1):31–6.

120. Yasuda T, Yamada Y, Hamatsu F, Hamagami T. A call triage support system for emergency medical service using multiple random forests. IEEJ Transactions on Electrical and Electronic Engineering. 2017;12:S67–S73.

121. Liu N, Koh ZX, Chua EC, Tan LM, Lin Z, Mirza B, et al. Risk scoring for prediction of acute cardiac complications from imbalanced clinical data. IEEE J Biomed Health Inform. 2014;18(6):1894–902.

122. Liu N, Sakamoto JT, Cao J, Koh ZX, Ho AFW, Lin Z, et al. Ensemble-Based Risk Scoring with Extreme Learning Machine for Prediction of Adverse Cardiac Events. Cognitive Computation. 2017;9(4):545–54.

123. Liu N, Guo D, Koh ZX, Ho AFW, Xie F, Tagami T, et al. Heart rate n-variability (HRnV) and its application to risk stratification of chest pain patients in the emergency department. BMC Cardiovasc Disord. 2020;20(1):168.

124. Topol EJ. Welcoming new guidelines for AI clinical research. Nat Med. 2020;26(9):1318–20.

125. Vasey B, Nagendran M, Campbell B, Clifton DA, Collins GS, Denaxas S, et al. Reporting guideline for the early-stage clinical evaluation of decision support systems driven by artificial intelligence: DECIDE-AI. Nat Med. 2022;28(5):924–33.

126. Acosta JN, Falcone GJ, Rajpurkar P, Topol EJ. Multimodal biomedical AI. Nat Med. 2022;28(9):1773–84.

127. Botker MT, Jacobsen L, Rudolph SS, Knudsen L. The role of point of care ultrasound in prehospital critical care: a systematic review. Scand J Trauma Resusc Emerg Med. 2018;26(1):51.

128. Kettner M, Helwig SA, Ragoschke-Schumm A, Schwindling L, Roumia S, Keller I, et al. Prehospital Computed Tomography Angiography in Acute Stroke Management. Cerebrovasc Dis. 2017;44(5-6):338–43.

129. Dewar A, Lowe D, McPhail D, Clegg G. The use of body-worn cameras in pre-hospital resuscitation. Br Paramed J. 2019;4(2):4–9.

130. Liu NT, Holcomb JB, Wade CE, Darrah MI, Salinas J. Data quality of a wearable vital signs monitor in the pre-hospital and emergency departments for enhancing prediction of needs for life-saving interventions in trauma patients. J Med Eng Technol. 2015;39(6):316–21.

131. Noorian AR, Bahr Hosseini M, Avila G, Gerardi R, Andrle A-F, Su M, et al. Use of Wearable Technology in Remote Evaluation of Acute Stroke Patients: Feasibility and Reliability of a Google Glass-Based Device. Journal of Stroke and Cerebrovascular Diseases. 2019;28(10):104258.

132. Markus AF, Kors JA, Rijnbeek PR. The role of explainability in creating trustworthy artificial intelligence for health care: A comprehensive survey of the terminology, design choices, and evaluation strategies. Journal of Biomedical Informatics. 2021;113:103655.

133. Duran JM, Jongsma KR. Who is afraid of black box algorithms? On the epistemological and ethical basis of trust in medical AI. J Med Ethics. 2021.

134. Volovici V, Syn NL, Ercole A, Zhao JJ, Liu N. Steps to avoid overuse and misuse of machine learning in clinical research. Nature Medicine. 2022;28(10):1996–9.

135. Loh HW, Ooi CP, Seoni S, Barua PD, Molinari F, Acharya UR. Application of explainable artificial intelligence for healthcare: A systematic review of the last decade (2011–2022). Computer Methods and Programs in Biomedicine. 2022;226:107161.

136. Ghassemi M, Oakden-Rayner L, Beam AL. The false hope of current approaches to explainable artificial intelligence in health care. Lancet Digit Health. 2021;3(11):e745–e50.

